# Optimizing Exomiser to prioritize candidate genes underlying monogenic forms of common disease

**DOI:** 10.64898/2026.11.20.26368494

**Authors:** Jana Petrovska, Isabel Rubio-Aliaga, Dorian T’Sjoen, Judith Van Wynsberghe, Olivier Vanakker, Johannes Münch, Elena M. Cabello, Olivier Bonny, Carsten A. Wagner, Ruxandra Bachmann-Gagescu

## Abstract

**Background:** The majority of rare diseases are caused by pathogenic variants in a single gene with Mendelian inheritance. In contrast, common diseases typically have complex genetic etiology, ranging from a cumulative impact of many common genetic variants with small individual effects, to fewer rare variants with larger effects, with monogenic forms at the rare end of the spectrum. While unknown prevalence of the monogenic fraction of common diseases and incomplete penetrance make identification of such monogenic forms of common diseases particularly challenging, their diagnosis can be essential for precision medicine. Automated rare variant prioritization tools, such as Exomiser, aid whole exome sequencing-based rare genetic disease diagnostics. However, Exomiser’s utility has not been systematically assessed in the context of monogenic forms of common diseases prior to this study.

**Methods:** We evaluated and optimized Exomiser for prioritization of rare pathogenic variants in common disease using a large kidney stone disease (KSD) cohort (701 cases, 200 controls). First, we benchmarked Exomiser’s performance across different settings for KSD “gold standard” variants in established KSD genes with Mendelian inheritance. Next, we utilized the optimal pipeline for discovery of novel KSD genes and compared Exomiser’s findings with results from gene-level rare variant enrichment analyses in an extended in-house cohort and the UK Biobank.

**Results:** With the optimal approach, Exomiser ranked within the top five rankings nearly all (94%, *N* = 48) previously defined “gold standard” KSD gene-participant pairs from an established KSD gene panel. Using this tool, we additionally nominated 62 novel genes predominantly involved in ion homeostasis, which are now available to the scientific community as candidate genes for KSD. Out of these genes, orthogonal approaches also support *ABCC6* as a strong KSD risk gene, in line with previous research, providing a proof-of-principle supporting the utility of Exomiser in the context of common disease.

**Conclusions:** This study serves as an example for automated variant prioritization in the context of monogenic forms of common diseases, which is a key step for precision medicine.

## Introduction

Genomics research in the last decades has greatly contributed to novel disease-gene discovery in both rare and common diseases. More than 70% of all rare diseases are considered to have a genetic origin with Mendelian inheritance, where disease results from mutation in a single gene (1). Identification of the causal gene responsible for disease now typically relies on whole exome sequencing (WES), which identifies a large number of genetic variants in any individual. Making the correct diagnosis then relies on properly prioritizing the most likely causative variant, depending on information from the rapidly growing resources for standardised disease and phenotype definition, such as Human Phenotype Ontology (HPO) (2), population-level genetic variant frequency estimation (GnomAD (3)), gene-to-phenotype mapping databases such as Online Mendelian Inheritance in Man (OMIM) (4,5) and in silico variant pathogenicity prediction algorithms (e.g., AlphaMissense (6), SpliceAI (7), CADD (8), etc.). To accelerate and improve this process, tools such as Exomiser, AI-MARRVEL, DeepRare, etc., that aggregate data from these resources, have been developed for rare-variant prioritization in Mendelian diseases (9–11). Such tools enable us to systematically and efficiently investigate variants in an exome-wide manner, look at a broader phenotypic context and rapidly update the search to incorporate novel discoveries (9,12,13).

In contrast to rare diseases, common diseases typically have a complex genetic etiology, with a cumulative impact of many common genetic variants with a relatively small individual effect. However, a subset of individuals with common diseases harbor one or a few rare variants with intermediate-to-large size effects, with monogenic forms at the rare end of the disease spectrum. These rarer monogenic forms can be phenotypically indistinguishable from common complex forms, as found across disease areas such as monogenic cancers (14), cardiovascular disease (15), neurodegenerative disorders such as Parkinson’s disease (16) and dementia (17), diabetes (18), inflammatory bowel disease (IBD) (19), chronic kidney disease (CKD) (20) or kidney stone disease (KSD) (21). Thus, the same disease can be caused either by pathogenic variants in a single gene in a monogenic form, or more frequently by the combined effects of multiple low-impact variants and environmental factors, whereby this does not represent a clear-cut dichotomy, but rather a spectrum. Near the rare end of this spectrum are genetic variants carrying strong risk, yet still needing additional genetic and/or environmental factors to reach the disease threshold, a situation often referred to as “monogenic with incomplete penetrance”.

Identifying monogenic forms of common diseases poses unique challenges. When aiming to identify causative gene variants with Mendelian inheritance in the context of rare diseases, all variants with high allele frequency in the population can be excluded given the link between disease prevalence and disease-allele frequency. Hence, variants with allele frequencies above 1% are typically excluded in the first steps of the analysis. In contrast, in common diseases the disease prevalence is high and the fraction of monogenic causes is typically unknown, so it is unclear where to place a threshold for allele frequency cut-off, or how to prioritize the more likely-to-be-diagnosed individuals (22,23). This is further complicated by the fact that monogenic forms of common diseases often have an incomplete penetrance (24,25). Despite these significant challenges, rare variant prioritization in the context of monogenic forms of common diseases is a key step towards better precision medicine. Certain monogenic forms of common diseases are well-established, and some already actionable, i.e., associated with changes to clinical management options for the patient (26). However, many more likely remain unidentified. In this context of common diseases, it appears particularly important to apply high throughput automated approaches to identify possible monogenic forms.

The most widely-adopted free, open-source tool for automated rare variant prioritization is Exomiser (27), with proven utility across Mendelian diseases (28–30). However, it has barely been used in the context of common diseases so far; once in the context of hereditary breast and ovarian cancer (31) and once for cardiomyopathies (32). While this tool has been recently optimized for use in the context of rare diseases (33), its performance when applied for capturing monogenic forms of common diseases has not been systematically evaluated.

KSD represents a very good model for systematic evaluation of Exomiser’s performance for identification of monogenic forms of common disease. KSD has a world-wide average lifetime prevalence of ∼10% and a high recurrence rate (34). It is caused by a complex interaction of genetic, environmental and dietary contributions (35). Genome wide association studies (GWAS) have found KSD-associated common variants at dozens of loci (36,37) and gene-level analysis has identified rare variant burden in *SLC34A3* as an additional KSD contributor (38), underlying the complex genetic architecture of this disease. Monogenic forms of KSD are well established with defined gene panels used in diagnostic practice (e.g. 35 high evidence, i.e., “green” genes for nephrocalcinosis or nephrolithiasis in the Genomics England PanelApp, v.6.3 (39)). Recently, we redefined the clinical relevance of genetic variants in a panel of such established KSD genes, by combining carefully curated genetic data with deep biochemical phenotyping in the large Swiss Kidney Stone Cohort (SKSC) (21). This integration of biochemical data impacted the interpretation of the clinical significance of genetic variants, resulting in 6.8% of adult kidney stone formers (KSF) carrying strong KSD risk variants, i.e., being at the “monogenic with incomplete penetrance” end of the spectrum. Taking these results as a “gold standard” variant dataset provided us with a unique opportunity to estimate Exomiser’s performance in the context of a common disease and subsequently use it for novel gene discovery.

We here present an optimization pipeline where we first evaluated and optimized Exomiser’s performance for rare variant prioritization in a common disease, KSD, across different settings by looking at gold standard variants in established KSD genes. We next utilized the optimized pipeline for novel gene discovery and compared Exomiser’s findings with results from gene-level rare variant enrichment analyses which aggregate (rare) variants within a gene across participants, as a more common method used in the context of novel common disease gene identification. Therefore, this work demonstrates how a tool with wide-spread use for automated genetic variant prioritization in rare diseases can be optimized and applied in the context of monogenic forms of a common disease.

## Methods

### SKSC study cohort

We used data from 716 adult KSF and 201 non-kidney stone formers (NKSF) from the SKSC, a multicenter longitudinal observational study. KSFs had either recurrent kidney stones or a single stone occurrence and at least one additional kidney stone (KS) -forming risk factor. Absence of KS was confirmed in the NKSF by computed tomography (CT). The precise study design and further participant characteristics have been described in more detail elsewhere (21,40).

### WES data acquisition and preprocessing

DNA was extracted from peripheral blood and WES was performed using the IDT xGen Exome Research Panel v2 (Integrated DNA Technologies, Coralville, Iowa, USA) on NovaSeq 6000 system (Illumina Inc.). Alignment and variant calling were done using DRAGEN Bio-IT Platform V3.9 (Illumina Inc.). The average coverage was 202. Exomes with a coverage < 100 were repeated. The average duplicate rate was 7.4, the average Q30 bases (having base-calling accuracy of 99.9%) was 92.72, and the average Ti/Tv ratio was 2.29. For more information please see (21).

Individuals with missing demographic information, mismatch between self-reported gender and WES-derived sex information, and genetic duplicate samples based on a WES kinship analysis were excluded from the study, resulting in 701 KSF and 200 NKSF remaining for downstream analyses.

### Data analyses

The data analysis workflow is depicted in Figure 1.

**Fig. 1.**
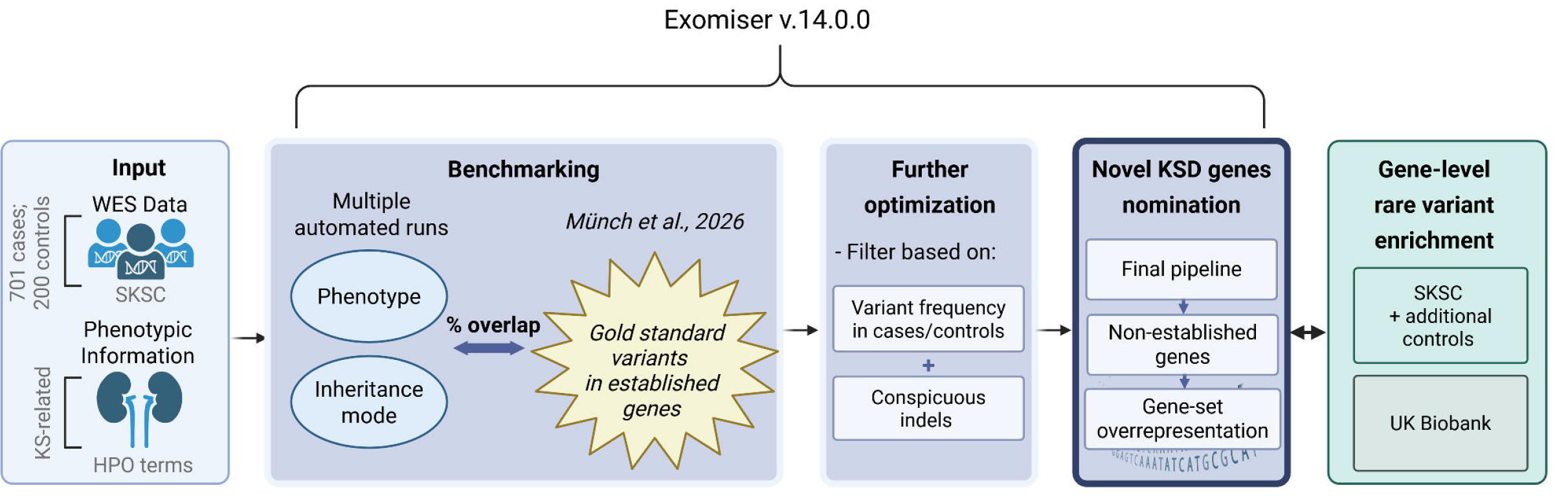
Workflow. An overview of the data analyses workflow. The input (SKSC WES data and a phenotypic list) is depicted in light blue; Exomiser-related steps are depicted in dark blue. The additional approach, i.e. gene-level rare variant enrichment analysis in-house (SKSC + additional controls) and in the UK Biobank is depicted in green. Created in BioRender. Petrovska, J. (2026) https://BioRender.com/c6mzu2h Abbreviations: WES, Whole Exome Sequencing; SKSC, Swiss Kidney Stone Cohort; KS, Kidney Stone; HPO, Human Phenotype Ontology.

### Exomiser

We used Exomiser v.14.0.0 for automated variant prioritization. This tool has been described in detail elsewhere (27,33). Briefly, it calculates gene-level variant and phenotype scores by incorporating state-of-the-art annotations and prediction algorithms. Next, it combines these scores producing a ranked list of protein-coding variants and genes across different inheritance modes, per individual (rank 1 = the most highly prioritized, 2 = second highest, etc.).

### Gene-set overrepresentation

We used the R package clusterProfiler (v4.20.0) (41) to identify gene sets in which our newly nominated KSD genes are overrepresented. Gene Ontology (GO): Biological Processes gene sets were considered. To remove redundancy from the relevant gene sets, we filtered based on semantic similarities using the R package simplifyEnrichment (42), v.2.6.0 (cutoff = 0.5), keeping only the most representative terms. We used a false discovery rate (FDR)-corrected p-value cutoff of 0.05 to determine statistical significance.

### Gene-level rare variant enrichment analyses

Gene-level analysis was performed with MAGMA v.1.10 (43). We applied the “multi=all” model which combines the resulting gene-level p-values of several base models into an aggregate p-value, to capture a wider range of different genetic architectures and have more even distribution of statistical power. For statistical power increase, we used WES data from additional in-house controls, acquired and preprocessed in an identical manner and in the same facility as the SKSC data. Ancestry was analyzed based on the genetic data in the overall sample (SKSC + additional controls) using the R package EthSEQ (v.3.0.2) (44) in 2D space. The majority of the overall sample was of European descent (Suppl. Figure 1), and non-European individuals were excluded from the gene-level analysis. We also excluded related individuals (up to second-degree), based on KING (45) v.2.3.1 robust kinship estimator, and a further check with PLINK (46) v.1.9 identity by descent (IBD) analysis. The final sample used in the gene-level analysis consisted of 638 cases and 621 controls, with a power > 0.80 for an odds ratio (OR) of 1.4 or higher, when testing a single gene (alpha = 0.05).

Ensembl Variant Effect Predictor (VEP), v115.1 (GRCh37/hg19) was used for variant annotation. We restricted the analysis to genetic variants with high or moderate impact according to VEP, minor allele frequency (MAF) < 0.02 in our sample and in the general population (based on the GnomAD v4.1 database), and Combined Annotation Dependent Depletion (CADD) PHRED-like scaled score >= 20, predicting the variant to be in the top 1% most deleterious possible substitutions in the human genome.

The first two principal components from the ancestry analysis were used as covariates in the gene-level analysis. We derived biological sex estimation based on the WES data and used it as an additional covariate.

Furthermore, we utilized gene-level association summary statistics from the UK Biobank sample, provided on the AstraZeneca PheWAS Portal: a public repository of gene-phenotype associations (47). We selected findings for “calculus of kidney”, phenotype code “41202#N200#”, resulting in 3,980 cases and 257,139 controls. Of note, the KS phenotype here is first KS occurrence, differing from the SKSC, where it is >1 KS occurrence or 1 KS occurrence + risk factor (see section “Study cohort”). We considered findings based on the following statistical models: “flexdmg”, “ptv5pcnt”, “ptvraredmg”, “raredmg”, and filtered for European ancestry only, for compatibility with the SKSC gene-level enrichment analysis.

### *ABCC6* cohort

Sequencing of the coding regions and exon-intron boundaries and multiplex ligation-dependent probe amplification (MLPA) of the *ABCC6* gene was performed at the Center for Medical Genetics Ghent, Belgium, as previously described (48). There were 222 heterozygous carriers of *ABCC6* variants in total, 159 of which carried (likely) pathogenic variants. For 96 of the heterozygous carriers of pathogenic variants inorganic pyrophosphate (PPi) levels were measured as previously described (49). Among the 159 carriers with pathogenic variants, 140 underwent ultrasonography and medical records were reviewed for systematic questioning regarding a history of nephrolithiasis. A composite renal calcification outcome was subsequently defined as the presence of either sonographically detected renal calcifications, a clinical history of nephrolithiasis, or both. Statistical analysis was performed in GraphPad Prism (version 10.6.1). For additional details on the methodology for the *ABCC6* cohort see supplementary material.

## Results

### Benchmarking and curation

As a first step, we investigated to what extent Exomiser prioritizes the rare variants in established KSD genes identified in our previous work through manual pathogenicity classification using the American College of Medical Genetics and Genomics/Association for Molecular Pathology (ACMG/AMP) criteria by an expert clinician combined with deep biochemical phenotyping (21). These variants served as our “gold standard” for benchmarking Exomiser performance in the context of kidney stones, serving as a model for common diseases. Of note, among these gold standard variants, some variants were deemed as causing “monogenic KSD” or as representing a “strong risk factor” for KSD in our previous work based on compelling biochemical evidence even when not aligned with the mode of inheritance previously reported in OMIM. Hence, heterozygous pathogenic or likely pathogenic variants were deemed (partially) causal in some genes such as *SLC34A3* despite being listed in OMIM only with recessive inheritance.

Taking advantage of this highly curated set of gold standard variants, we applied Exomiser on the WES data of the SKSC to test and optimize its performance for this common disease. For each variant in each individual, Exomiser provides a phenotype score (depending on the phenotypes of interest indicated for that individual) and a variant score (predicting the deleteriousness of the variant). These scores range from 0 to 1 and are combined to create a single gene score, with higher scores contributing to higher prioritization ranking. We initially executed Exomiser v.14.0.0 with default settings and “nephrolithiasis” (kidney stones) (HP:0000787) as the phenotype of interest. Variants not passing standard quality control were disregarded. We considered genes with a phenotype score > 0.2 and a variant score > 0.8 to be of interest. The variant score considers the minor allele frequency (MAF) in the general population (the highest MAF from multiple ancestries taken into account), and excludes variants with MAF above a predefined threshold. It also includes algorithms for in silico variant pathogenicity prediction, incorporating the most severe prediction in the variant score, when multiple algorithms are considered. The population MAF threshold, the pathogenicity prediction algorithms used, as well as other metrics, can be modified by the user.

For SKSC individuals with a gold standard variant in an established KSD gene, we looked at the Exomiser ranking of the respective gene based on its phenotype and variant scoring. As part of the variant-level scoring, we used the following algorithms for in silico variant pathogenicity prediction: MVP, REVEL, AlphaMissense, and SpliceAI, as recommended by a previous optimization study for rare diseases (33). The MAF cutoff for disregarding variants which are too common in the general population to be of interest remained at the default setting of 2% for compound heterozygous models and was raised from 0.1% to 1% for autosomal dominant and recessive - homozygous alternative models. The exact prevalence of monogenic KSD in the general population remains uncertain. However, we considered 1% as an approximate estimate, considering KSD general population prevalence of ∼ 10% (34) and ∼ 10% of KSD to be monogenic. The latter estimate is based on our previous study with 6.7% of the SKSC identified as monogenic (21), allowing for some additional missed cases. Furthermore, variants ranking top 1 in > 10% of the SKSC sample were disregarded, given the high false positive likelihood. Everything ranking below them was ranked up +1 to compensate.

We estimated the performance of each Exomiser run by calculating the proportion of gold standard variants in established genes (21), using increasing rank thresholds from 1 to 30. We did not consider variants with rank above 30 since this was adding too much noise to the results, in line with Exomiser optimization research in rare diseases (33). The variant prioritization was considered to be successful if the previously established gene was prioritized by Exomiser in the same individual, regardless of the specific variant and mode of inheritance (MOI), i.e., gene-level success.

In our initial analysis, the previously identified gold standard variant was identified by Exomiser within the top 30 rankings in 63 % (*N* = 32) of carriers. To optimize Exomiser’s performance, we successively re-ran the analyses with the following setting alterations:

#### 1) Expanding the list of phenotypes

Multiple phenotypes (HPO codes) can be provided to Exomiser for the same individual and broadening the search by extending the phenotype list can be beneficial in certain cases. Therefore, an expert nephrologist provided a list of terms biologically linked to nephrolithiasis (e.g., hypercalciuria, hypocitraturia, etc.) which were then mapped onto HPO codes (Suppl. Table 2), *N* = 31. When multiple HPO codes were available, the most general one was selected.

#### 2) Disregarding the mode of inheritance

Exomiser uses OMIM inheritance mode information by checking if a patient’s candidate genotype is consistent with the reported disease inheritance pattern and halves the phenotype score in case of inconsistency. As previously mentioned, the mode of inheritance for our “gold standard” variants did not always align with what is reported in OMIM for the respective established gene. This is due to the “gold standard” utilizing an additional level of information - blood and urine biochemistry data. For example, *SLC34A3* is listed only under autosomal recessive inheritance in OMIM, while it was identified as autosomal dominant in our “gold standard” list and confirmed by other cohorts (50,51). Given the very recent publication of this list, it has not yet been incorporated in OMIM. Therefore, we reran Exomiser disregarding the OMIM inheritance mode contribution to the scoring.

As indicated above, when focusing on the first 30 ranked genes per participant and selecting “nephrolithiasis” as the only phenotype Exomiser found the previously established variants in 63% (*N* = 32) of cases. When focusing only on the first one or the first five top-ranked genes with default values, we identified 59% (*N* = 30), and 63% (*N* = 32) carriers, respectively (Figure 2A). With the optimized pipeline, extending the phenotype list and disregarding inheritance mode, 94% (*N* = 48) of the variants were identified within the top 5 ranks. Including variants further down the list did not substantially improve performance beyond rank 5. Only 1 additional carrier was identified when extending to rank <= 30, reaching a total of 96% (49 out of 51 carriers prioritized).

**Fig. 2.**
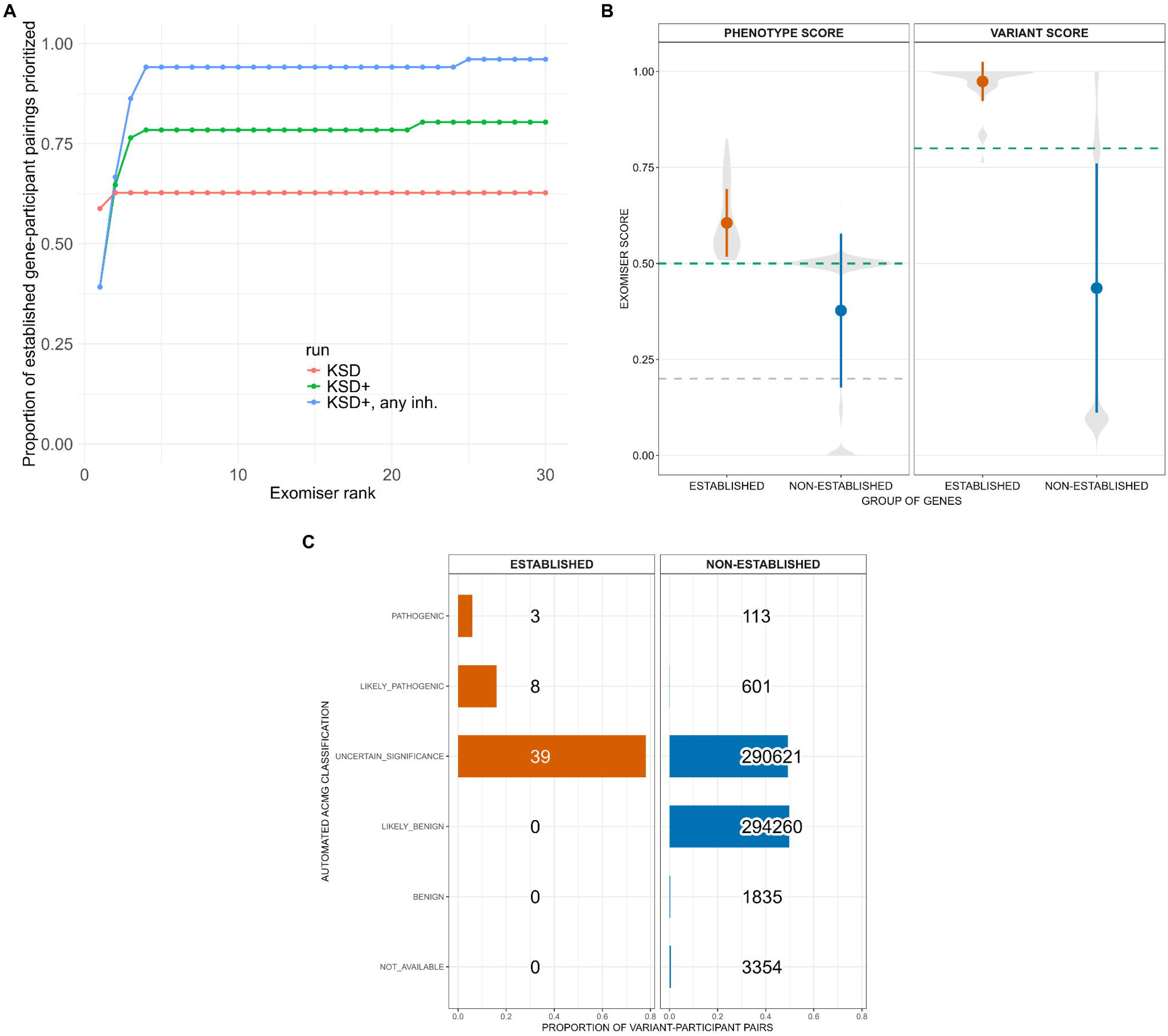
Benchmarking Exomiser in the context of kidney stone disease. (A) Proportion of carriers of gold standard monogenic variants in established genes which were also identified by Exomiser within the top X rankings (X-axis). Results from analyses with different Exomiser settings are depicted in different colors: “KSD” (red) = KSD as the only phenotype; “KSD+” (green) = KSD and additional related phenotypes included (see Suppl. Table 2); “KSD+, any inh.” (blue) = KSD and related phenotypes, and OMIM inheritance removed from the scoring. (B) Violin plots depicting a comparison of the distribution of Exomiser scores between gold standard variants in established KSD genes and all other variants in all other genes (=” non-established”) for variant and phenotype-level scores, respectively. The initial cutoff score for results filtering is depicted as a gray dashed line, and the cutoff deemed as optimal as a green dashed line (in case they overlap, only the green line is seen). The error bars represent the standard deviation from the mean. (C) Overview of Exomiser’s automated variant classification based on ACMG criteria per variant-participant pairing within “non-established” (in blue) and established genes (in red), respectively. The numbers indicate the number of variants in each ACMG category. As expected, for non-established genes, where all variants were considered for ranking, all ACMG categories (P, LP, VUS, LB, B) are represented while for the established genes the variants preselected for disease relevance (gold standard) do not include LB/B variants. Note that the majority of variants among established genes are classified as VUS even though they represent the gold standard variants used for benchmarking, illustrating how the addition of detailed phenotypic data (in this case biochemical information for each participant) improves ACMG classification compared to the Exomiser automated method.

The two unidentified gene-participant pairings were for the genes *SLC4A1* and *SLC34A3*, respectively. In the case of *SLC4A1*, this was due to the variant (c.713A>T) having a GnomAD MAF slightly above the set threshold. The missed variant in *SLC34A3* (c.1058G>T) was filtered out due to a variant score of 0.76. Given that this was the only “gold standard” variant with variant score below our set threshold of 0.8 (Figure 2B), we decided not to lower the threshold to avoid introducing noise when extending to novel genes.

As all gold standard variants had a phenotype score > 0.5 (Figure 2B), we decided to increase our initially set threshold of 0.2 to 0.5 for this metric. The majority of the gold standard variants were assigned as variants of unknown significance (VUS) by the automated Exomiser assignment (Figure 2C), indicating that the addition of clinical data (in this case biochemical data relevant for the analyzed genes) improves variant classification as expected.

### Nominating candidate novel kidney stone genes

After having used the previous gold standard dataset to optimize Exomiser in this context and ensuring that it successfully prioritizes the majority of manually found established KSD genes with clinically relevant variants, we next moved towards nominating novel candidate genes for monogenic forms of KSD among genes not associated with KSD yet. Considering our large sample size, we focused only on variants with rank 1, for noise reduction.

We used the following filtering criteria when evaluating putative novel KSD genes: a) Variants within a gene must be present in at least two cases.; b) A variant can be present in no more than 1% of cases or controls.; c) The odds ratio for a given gene is > 0.25 (to remove genes where the odds of carrying a variant within that gene are substantially higher in controls than in cases: this filtering step removes obvious negatives but keeps genes with, e.g., 1 carrier in cases and 1 in controls to avoid being too stringent, as controls may have a relevant variant with incomplete penetrance and/or develop a KS later in life).; d) The variant carrier was filtered out if the individual was also a carrier of a gold standard variant in an established KSD gene (i.e. already had a monogenic cause identified); e) The variant should not be classified as (likely) benign by Exomiser’s automated ACMG criteria assignment.

Using this optimized pipeline, we identified 62 genes as novel candidates for KSD. A list of all identified genes can be found in Suppl. Table 3, and more detailed information regarding the variants identified (*N _total_* = 176: 159 VUS; 16 LP; 1 P, as assigned by Exomiser) and their carriers (including urine and blood biochemistry) is available per gene from our online browser: https://sksc-variant-and-biochemistry.shinyapps.io/exomiser-candidates/. Based on gene-set overrepresentation analysis, we found these genes to be predominantly involved in purine nucleotide metabolic process (GO:0006163) and in monoatomic cation homeostasis (GO:0055080) (Figure 3A). 49 of the 62 genes nominated by Exomiser already have an OMIM-recorded disease association while 13 have not yet been described as monogenic disease genes. Among the 49 candidate genes known to be associated with other non-KSD diseases, we identified *ATP7B, FGF23, GNA11* and *PHEX,* which have already been associated to some extent with KSD and are already included in some, but not all, KSD genetic testing panels (52). *FGF23* and *GNA11* for example are also included in the Genomics England PanelApp for nephrocalcinosis or nephrolithiasis as candidate “red” genes, i.e., genes with low level of evidence - supported by a single source or only preliminary data (v6.3; https://panelapp.genomicsengland.co.uk/panels/149/ (39)).

**Fig. 3.**
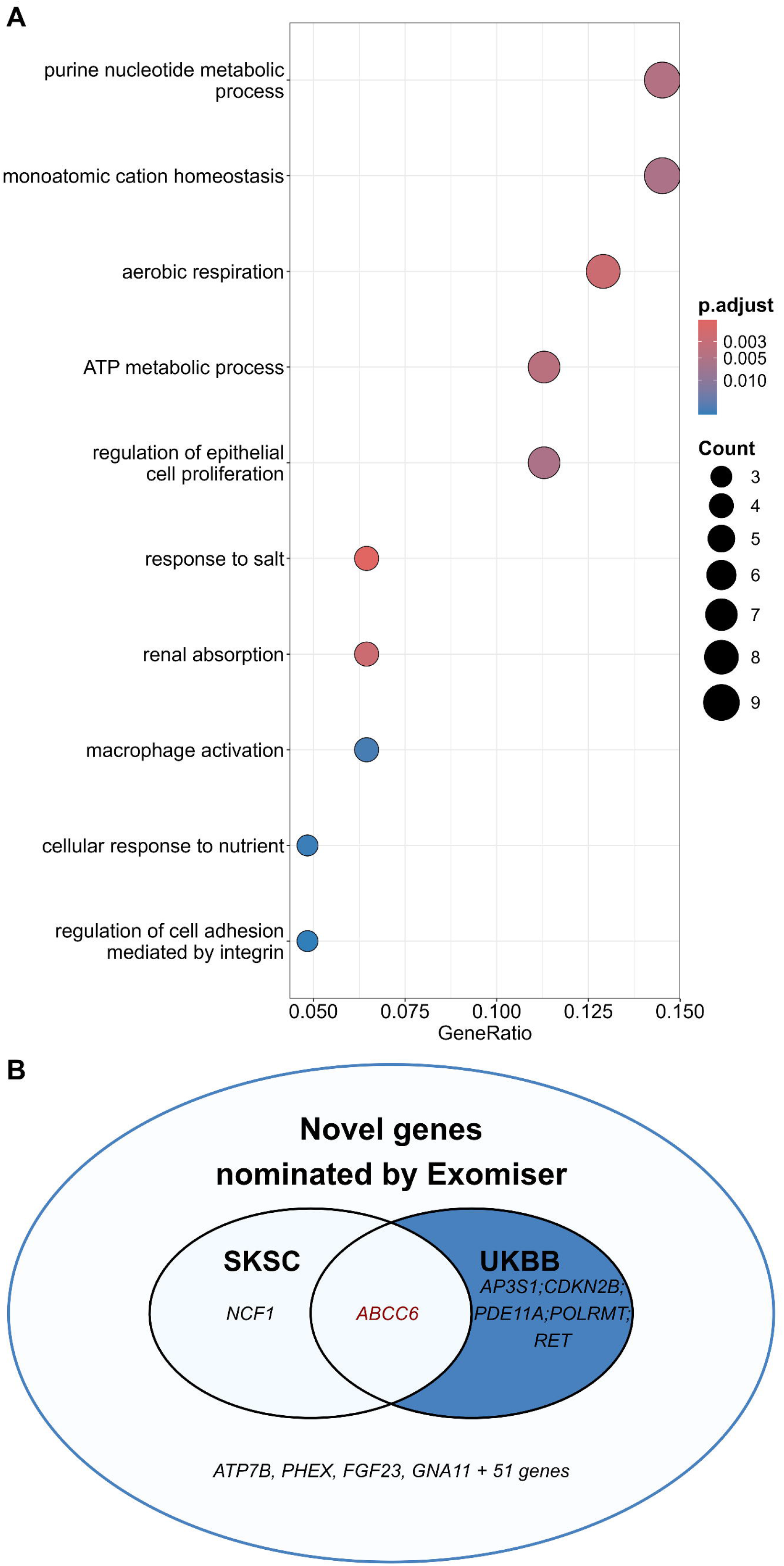
Gene-set overrepresentation for novel nominated genes and comparison with rare variant enrichment analyses. (A) Top 10 gene sets with overrepresented novel nominated genes. (B) Venn diagram comparing the novel nominated genes (ranked top 1 by Exomiser) that also have nominally significant (*p* < 0.05) gene-level rare variant enrichment for KSD in the extended Swiss Kidney Stone Cohort (SKSC) and in the UK Biobank (UKBB) for “calculus of the kidney”.

Similarly, *ATP7B* or *CFTR* cause distinct disorders (Wilson disease and cystic fibrosis respectively), where some evidence of association with kidney stones has been previously reported (53,54). This supports the utility of our approach, as these genes, while clearly associated with other monogenic conditions, appear to also be involved in KSD, probably through secondary effects such as dysregulation of calcium and phosphate metabolism by *PHEX*, *GNA11* or *FGF23* mutations.

Two of the newly nominated genes, namely *MUC2* and *MUC20*, are on Exomiser’s black list due to having a high number of variants called in healthy individuals. Since having a kidney stone is not a typical exclusion criterium when recruiting healthy control participants in most studies and since there is already some evidence for the involvement of another mucin gene (*MUC1)* in KSD (55,56), we decided not to exclude these genes from the nominated list.

To further explore the plausibility of an association between KSD and the candidate genes nominated by Exomiser, we next investigated if we could identify some of these novel nominated genes through other methods.

### Comparing Exomiser-nominated genes with gene-level rare variant enrichment

As an orthogonal approach for identification of novel candidate genes for monogenic KSD, we performed a gene-level rare variant enrichment analysis to identify KSD-candidate genes in the SKSC. To improve statistical power, we increased the number of control samples by including WES data from in-house controls sequenced in the same manner (*N* = 621). Unfortunately, no gene reached genome-wide significance for KS association in the gene-level analysis (Suppl. Figure 2), which is not surprising given the limited sample size.

However, 2 out of the 62 newly nominated genes through Exomiser reached nominal significance in this gene-level rare variant enrichment analysis (*p* < 0.05): *NCF1* and *ABCC6* (Suppl. Table 4 and Suppl. Figure 3). Both of these are known disease genes, where bi-allelic pathogenic variants in *NCF1* cause chronic granulomatosis disease while bi-allelic pathogenic variants in *ABCC6* are responsible for pseudoxanthoma elasticum. Both disorders are inherited in a recessive manner. In pseudoxanthoma elasticum, *ABCC6* dysfunction leads to reduced ATP transport and decreased levels of the circulating calcification inhibitor PPi, which in turn causes abnormal tissue calcifications including formation of kidney stones in some patients (57).

In an independent approach, we also utilized the UK Biobank data for gene-level enrichment in individuals with KS. This identified 6 of the Exomiser-nominated genes that reached nominal significance (*p* < 0.05) (Suppl. Table 4), among which we found again *ABCC6*. An overview of the overlap of nominated genes that were found by the gene-level approaches (at least on a nominal level) is provided in Figure 3B. Of note, *ABCC6* was the only gene significantly associated with KS in both the SKSC and the UK Biobank sample (SKSC: *p* = 0.007; UK Biobank: *p* = 0.0005). We therefore focused on this gene for further exploration.

We next manually classified all rare (MAF < 0.01), exonic *ABCC6* variants in the SKSC according to the ACMG criteria. 80 individuals had an *ABCC6* variant, of which 31 harbored one likely pathogenic/pathogenic (LP/P) variant, 10 a likely benign/benign (LB/B) and 41 a variant of unknown significance (VUS). All but one individual were heterozygous for the LP/P *ABCC6* variants. Two individuals had two variants each, the first carrying a P and a VUS variant, and the second carrying a VUS and a B variant (whereby the phase cannot be established here based on WES data alone). There were 10 unique LP/P variants, reported in Table 1. Importantly, LP/P variants in *ABCC6* were significantly more prevalent in KSF than in CT-confirmed NKSF (OR = 8.9, *p* = 0.007). Two out of the ten variants of interest were ranked top 1 by Exomiser. All *ABCC6* variants, including VUS and LB/B variants are reported in Suppl. Table 6.

**Table 1.** LP/P *ABCC6* variants in SKSC.

| Variant ID | ACMG | HGVSc | HGVSp | Consequence | EXON | INTRON | GT | gnomADe MAF | CADD | MAC Case | MAC Control | Exomiser Rank |
|---|---|---|---|---|---|---|---|---|---|---|---|---|
| HIGH IMPACT |  |  |  |  |  |  |  |  |  |  |  |  |
| **16:16256935:G:A** | **P** | **NM_001171.6: c.3421C>T** | **NP_001162.5: p.(Arg1141Ter)** | **stop gained** | **24/31** | **-** | **0/1** | **1.7e-03** | **41.0** | **4** | **0** | **1<sup>1</sup>** |
| 16:16248510:AC:A | LP | NM_001171.6: c.4182del | NP_001162.5: p.(Lys1394AsnfsTer9) | frameshift | 29/31 | - | 0/1 | 3.2e-05 | 27.3 | 1 | 0 | 5<sup>2</sup> |
| 16:16308234:G:GC | LP | NM_001171.6: c.546dup | NP_001162.5: p.(Leu183AlafsTer13) | frameshift | 5/31 | - | 0/1 | 1.3e-05 | 24.6 | 1 | 0 | 2<sup>3</sup> |
| 16:16251667:C:T | P | NM_001171.6: c.3736-1G>A | - | splice acceptor | - | 26/30 | 0/1 | 1.6e-05 | 35.0 | 1 | 0 | 3<sup>4</sup> |
| MODERATE IMPACT |  |  |  |  |  |  |  |  |  |  |  |  |
| **16:16272807:C:T** | **LP** | **NM_001171.6: c.2263G>A** | **NP_001162.5: p.(Gly755Arg)** | **missense** | **18/31** | **-** | **1/1** | **2.1e-05** | **27.1** | **2** | **0** | **1<sup>1</sup>** |
| 16:16295863:T:C | LP | NM_001171.6: c.1171A>G | NP_001162.5: p.(Arg391Gly) | missense | 9/31 | - | 0/1 | 7.0e-03 | 21.0 | 17 | 1 | 142 |
| 16:16282764:A:G | P | NM_001171.6: c.1703T>C | NP_001162.5: p.(Phe568Ser) | missense | 13/31 | - | 0/1 | 4.7e-05 | 28.3 | 2 | 0 | 23 |
| 16:16248755:C:T | P | NM_001171.6: c.4016G>A | NP_001162.5: p.(Arg1339His) | missense | 28/31 | - | 0/1 | 1.9e-05 | 29.3 | 1 | 0 | 2<sup>5</sup> |
| 16:16248879:C:T | LP | NM_001171.6: c.3892G>A | NP_001162.5: p.(Val1298Ile) | missense | 28/31 | - | 0/1 | 2.6e-04 | 27.2 | 1 | 0 | 78 |
| 16:16248831:G:A | P | NM_001171.6: c.3940C>T | NP_001162.5: p.(Arg1314Trp) | missense | 28/31 | - | 0/1 | 7.9e-05 | 26.2 | 1 | 0 | 2<sup>6</sup> |

For LP/P *ABCC6* variants with MAF > 0.001 we additionally checked for a variant-level kidney stone association in the UK Biobank. This identified two such variants, both with significant kidney stone association (NM_001171.6: c.3421C>T: OR = 2.01, *p* = 0.0006; NM_001171.6: c.1171A>G: OR = 1.42, *p* = 0.002), Suppl. Table 7.

We next analyzed the clinical and biochemical data in a cohort of individuals with heterozygous LP/P *ABCC6* variants (*N*=159) from Belgium. 15.72% (25/159) of individuals had renal calcifications and/or clinical episode(s) of lithiasis. Since the precise prevalence of KSD and of renal calcifications is unknown in this population, it remains unclear whether this represents a significantly increased incidence. Analysis of the plasma PPi levels in heterozygous LP/P *ABCC6* variant carriers did not show a significant difference between individuals with or without a history of kidney stones (Suppl. Figure S4). Despite this, the genetic association between LP/P *ABCC6* variants identified through our Exomiser approach is supported by the gene-level rare variant enrichment analysis in the SKSC and in UK biobank, by the variant-level association in the UK biobank and by previous literature (57,58).

## Discussion

This study shows how a widely used tool for variant prioritization developed for rare diseases can be used for identifying monogenic forms of a common, complex disease. The availability of a large and carefully curated dataset, where genetic variant interpretation is supported by biochemical data, represents an ideal setting for benchmarking Exomiser in the context of a common disease. After optimizing the settings for our purpose by extending the phenotypic list and disregarding previously assigned inheritance modes, 94% of the “gold standard” gene-participant pairings were prioritized by Exomiser within the top 5 rankings, indicating good performance. Since inclusion of variants ranked below 5 did not substantially increase the performance, rank 5 may be an optimal cutoff in the context of (this) common disease, where carefully balancing the signal-to-noise ratio is particularly important due to the expectation that the majority of participants are not affected by monogenic forms of the disease.

We further extended this pipeline to utilize Exomiser for novel gene discovery and nominated 62 genes not previously considered causative for monogenic KSD as potentially relevant for kidney stones. Interestingly, several of these genes have already been tentatively associated with KSD (52), serving as a positive control for our approach and indicating that we can successfully identify relevant signals by “casting a wider net”. This is further supported by the orthogonal approach relying on gene-level enrichment analysis in the SKSC and the UK Biobank, which both identified the adenosine triphosphate (ATP) binding cassette subfamily C member 6 (*ABCC6*) as associated with KSD. *ABCC6* is a transmembrane protein predominantly expressed in the liver and the kidney (59).

Mutations in this gene are linked to low levels of plasma PPi, an inhibitor of calcium crystallization, and have been associated with two rare diseases: generalized arterial calcification of infancy (GACI) (60) and pseudoxanthoma elasticum (PXE) (61). PXE is an autosomal recessive multisystem rare genetic disorder characterized by elastic fiber calcification in the skin, the cardiovascular system, and the eyes, and has also been previously associated with an increased KSD risk (57,62–64). Heterozygous *ABCC6* mutations have also been reported in individuals with recurrent KSD (58) and linked to kidney stone occurrence as part of a large population copy number variation analysis (65). Therefore, this gene is indeed considered as a strong candidate risk factor for KSD and prioritizing it by an exome-wide automated search may be seen as a proof of principle for utilization of this methodology for identification of genes linked to monogenic forms of common diseases.

The lack of significant association between plasma PPi levels and history of kidney stones in an independent cohort of heterozygous *ABCC6* carriers reported in this study does not exclude an effect for heterozygous LP/P *ABCC6* variants acting as risk factors for KSD. The small number of individuals limits statistical power and it remains uncertain how much a single plasma PPi measurement is representative of tissue-relevant levels, e.g., urine or kidney levels. Given the converging genetic evidence for *ABCC6* involvement in KSD through Exomiser, rare variant enrichment analyses in the SKSC cohort and the UK Biobank, as well as previous reports, it appears likely that LP/P variants in this gene represent a strong risk factor, and place *ABCC6*-associated KSD towards the “monogenic with incomplete penetrance” end of the disease spectrum.

This study illustrates the potential utility of automated variant prioritization for the investigation of monogenic forms of common diseases. This approach can now also be applied to identify monogenic forms of other common diseases in large cohorts, where its performance should be tested. Indeed, some limitations must be taken into account, as is the case with all automated approaches but particularly in the context of common diseases. As with a lot of genetic data, a good gold standard dataset for benchmarking is hard to obtain, since it is often difficult to prove the pathogenicity of genetic variants without functional studies. In this study, we could take advantage of deep phenotypic information including biochemical data to support our interpretation of the effects of variants (21). Generating a reliable gold standard dataset for benchmarking can however represent a challenge in other disorders where biochemical data are not available or as relevant. Applying the Exomiser settings defined here without disorder-specific benchmarking may be of value but would require some positive controls in the dataset to evaluate the overall performance. A major challenge in this context is the uncertainty about the actual proportion of monogenic cases in various common diseases, which directly influences the MAF cut-offs for Exomiser. This is also linked to the fact that incomplete penetrance is frequent in monogenic cases of common diseases and that disease may manifest later in life. Consequently, controls may carry strong disease risk alleles. In this case, we estimated based on previous work that approximately 10% of KSD may be of or close to monogenic nature with a strong risk factor in one gene. Since the overall prevalence of KSD is around 10%, this allowed us to determine a MAF cut-off of 1% (10% of 10%). This would certainly need to be adapted to each disorder group based on current disease prevalence data.

One limitation of our study is that the majority of participants in the SKSC were of European ancestry. Therefore, how these findings translate to other ancestries remains to be systematically investigated in future studies. Moreover, here we only focused on coding variants using WES. Whole genome sequencing (WGS) and utilizing Genomiser (a variant prioritization tool including non-coding regions) has been suggested as a second step for WES-undiagnosed individuals in rare diseases (33). Given the difficulty in interpreting the significance of non-coding variants even in rare monogenic diseases, this may prove to be even more challenging in the context of rare monogenic forms of common disease, where the proportion of expected monogenic cases is unclear. Finally, while the use of Exomiser is well-established in the context of Mendelian diseases, it is not the only tool available. The choice for rare variant prioritization software is rapidly growing (10,11,66–68), and it remains to be seen how the performance of other automated tools will compare in the context of KSD and other common diseases.

In summary, we systematically investigated the use of automated variant prioritization with Exomiser in a large KSD cohort, as an example for a common disease with monogenic forms. With the optimal approach, Exomiser ranked within the top five rankings nearly all previously defined “gold standard” KSD gene-participant pairs from an established KSD gene panel. Using this tool, we additionally nominated novel genes which are now available to the scientific community for further investigation (https://sksc-variant-and-biochemistry.shinyapps.io/exomiser-candidates/). Using orthogonal approaches, we present *ABCC6* as an example of a strong KSD candidate gene, also in line with previous research. Therefore, this study may serve as an example of automated variant prioritization in the context of monogenic forms of common diseases which is a key step for precision medicine, given its potentially high clinical utility. In the case of KSD, we could previously show that having a rare genetic variant with a strong impact in KSD-related genes is predictive of kidney stone recurrence (21) which may impact the clinical management of affected individuals. With this study, we take a step further toward the successful diagnosis of monogenic forms of common diseases, with potential impact on disease prognosis and treatment.

## List of abbreviations

*ABCC6*: ATP binding cassette subfamily C member 6
ACMG: American College of Medical Genetics and Genomics
AMP: Association for Molecular Pathology
ATP: adenosine triphosphate
B: benign (ACMG category)
CADD: Combined Annotation Dependent Depletion
CKD: chronic kidney disease
CT: computed tomography
FDR: false discovery rate
GACI: generalized arterial calcification of infancy
GO: Gene Ontology
GWAS: genome wide association study
HHRH: hereditary hypophosphatemic rickets with hypercalciuria
HPO: Human Phenotype Ontology
IBD: inflammatory bowel disease
KS: kidney stone
KSD: kidney stone disease
KSF: kidney stone formers
LB: likely benign (ACMG category)
LP: likely pathogenic (ACMG category)
LP/P: likely pathogenic/pathogenic
MAF: minor allele frequency
MATK: Mendelian Analysis Toolkit
MLPA: multiplex ligation-dependent probe amplification
MOI: mode of inheritance
NKSF: non-kidney stone formers
OMIM: Online Mendelian Inheritance in Man
OR: odds ratio
P: pathogenic (ACMG category)
PPi: inorganic pyrophosphate
PXE: pseudoxanthoma elasticum
SKSC: Swiss Kidney Stone Cohort
UKBB: UK Biobank
VEP: Variant Effect Predictor (Ensembl)
VUS: variants of unknown significance
WES: whole exome sequencing
WGS: whole genome sequencing

## Declarations

### Ethics approval and consent to participate

Informed consent was given by all participants. The genetics of SKSC study was approved by the Cantonal Ethics Commission of the Canton of Zurich (BASEC 2021-00589). The *ABCC6* study was approved by the Ethical Committee of the Ghent University Hospital, and the tenets of the Declaration of Helsinki were followed.

### Availability of data and materials

The main datasets supporting the conclusions of this article are included within the article (and its additional files), and available from our genomic browser: https://sksc-variant-and-biochemistry.shinyapps.io/exomiser-candidates/. Any additional data available will be shared upon request to the corresponding authors (except for per-patient WES data for data protection reasons).

The gene-level and variant-level association summary statistics from the UK Biobank sample are freely accessible from the AstraZeneca PheWAS Portal (https://azphewas.com/).

The code used for this study is available from the following GitHub repository: https://github.com/PetJana/optimizing_exomiser_common_disease.

The Exomiser tool, along with the supporting resources, is freely available and accessible from the following GitHub repository: https://github.com/exomiser/Exomiser.

MAGMA - the software used for gene-level enrichment analysis is freely available, along with the supporting resources, from https://cncr.nl/research/magma/.

### Competing interests

The authors declare that they have no competing interests.

### Funding

National Center for Competence in Research NCCR Kidney.CH, funded by the Swiss National Science Foundation (183774) and the University of Zurich to OB, CAW, RBG. CAW was supported by a grant from the Swiss National Science Foundation (10007228).

### Authors’ contributions

Study design and conception J.P., R.B.-G., and C.A.W.; recruitment of participants C.A.W.,

O.B and O.V.; generation and interpretation of data J.P., C.A.W., R.B.-G., J.M., I.R.-A, E.M.C. D.T., J.V.W, O.V; manuscript writing J.P. and R.B.-G.; manuscript reviewing and editing all authors.

## Acknowledgements

We are very grateful to all study participants, as well as staff collecting the data, without whom this study would not have been possible.

## Notes

### Competing Interest Statement

The authors have declared no competing interest.

### Author Declarations

The Cantonal Ethics Commission of the Canton of Zurich (BASEC 2021-00589) gave ethical approval for the genetics of SKSC study. The Ethical Committee of the Ghent University Hospital gave ethical approval for the follow-up ABCC6 study and the tenets of the Declaration of Helsinki were followed.

