## Supplementary Information and Figures for "Optimizing Exomiser to prioritize candidate genes underlying monogenic forms of common disease"

**Supplementary Methods**

**Supplementary Figures**

**Suppl. Figure 1**: Genetic ancestry (SKSC + additional controls)

**Suppl. Figure 2**: Results from gene-level analysis with MAGMA for all genes

**Suppl. Figure 3**: Results from gene-level analysis with MAGMA for novel, i.e., non-established genes nominated by Exomiser

**Suppl. Figure 4:** Inorganic plasma pyrophosphate levels in heterozygous carriers of LP/P *ABCC6* variants

**Supplementary References**

### Supplementary methods

#### *ABCC6* cohort details and analysis

In total 222 heterozygous carriers were included through the PXE Reference Center of the Center of Medical Genetics (CMGG) of the Ghent University Hospital.

Sequencing of the coding regions and exon-intron boundaries of the PXE-related genes (*ABCC6, ENPP1, GGCX*) and multiplex ligation-dependent probe amplification (MLPA) of the *ABCC6* gene was performed at the Center for Medical Genetics Ghent as previously described (1). Variants were classified according to the guidelines published by Verschuere et al., using the most recent available evidence (2). Variants classified as variants of unknown significance and variants with incomplete penetrance, as previously described by Szeri et al. (3), were excluded. 159 individuals carried (likely) pathogenic variants.

For 96 heterozygous carriers of pathogenic variants, non-fasting blood samples were collected in four citrate BD Vacutainers (363,083 BD) and stored on ice. Blood samples were immediately centrifuged at 1000 x g for 10 minutes at 4°C. Plasma was pooled and redistributed into Centrisart I MWCO 300,000 Da tubes (Item number 13,279-E, Sartorius, Göttingen, Germany) and centrifuged at 2300 x g for 35 minutes at 4°C. Purified plasma was collected in 1.5ml Eppendorf tubes and stored at -80°C until PPi measurement. Plasma PPi content was measured using a two-step ATPase luminescence assay, as previously described by Van Gils et al. (4). Statistical analysis was performed in GraphPad Prism (version 10.6.1).

Among the 159 carriers with a class 4 or class 5 variant, 140 underwent ultrasonography of the kidneys as part of the clinical work-up following identification of their carrier status. This work-up was performed in accordance with the previously described screening approach for heterozygous *ABCC6* carriers (5), which includes ultrasonographic evaluation of the abdominal organs. The original ultrasound reports were retrospectively retrieved from the participants' electronic medical records. The presence or absence of renal calcifications was recorded as reported by the examining radiologist. No central review or retrospective reinterpretation of the ultrasound images was performed.

In addition, the medical records of all carriers were reviewed for systematic questioning regarding a history of nephrolithiasis. A history of nephrolithiasis was considered present when kidney stones had been reported by the participant or documented in the medical record. When available, nephrolithiasis was classified as a single or recurrent event. Sonographically detected renal calcifications and a clinical history of nephrolithiasis were initially recorded as separate outcomes. A composite renal calcification outcome was subsequently defined as the presence of either sonographically detected renal calcifications, a clinical history of nephrolithiasis, or both.

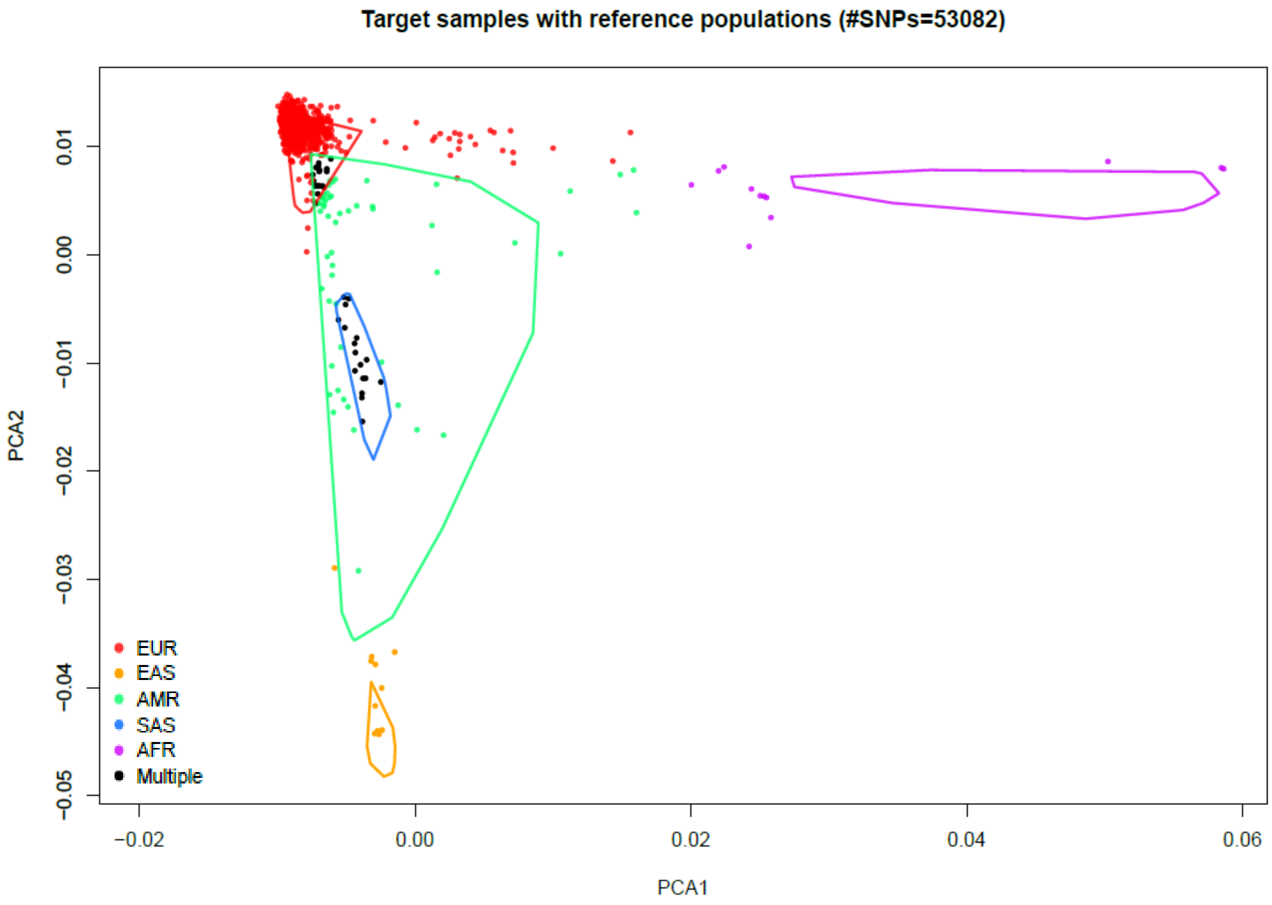


**Suppl. Figure 1: Genetic ancestry (SKSC + additional controls)**

Ancestry was analyzed based on the genetic data using the R package EthSEQ (v.3.0.2), in 2D space.

Abbreviations: SNPs, Single Nucleotide Polymorphisms, PCA1, The first component from the Principal Component Analysis (PCA), PCA2, The second component from the PCA, AFR, African, AMR, Ad Mixed American, EAS, East Asian, EUR, European, SAS, South Asian.

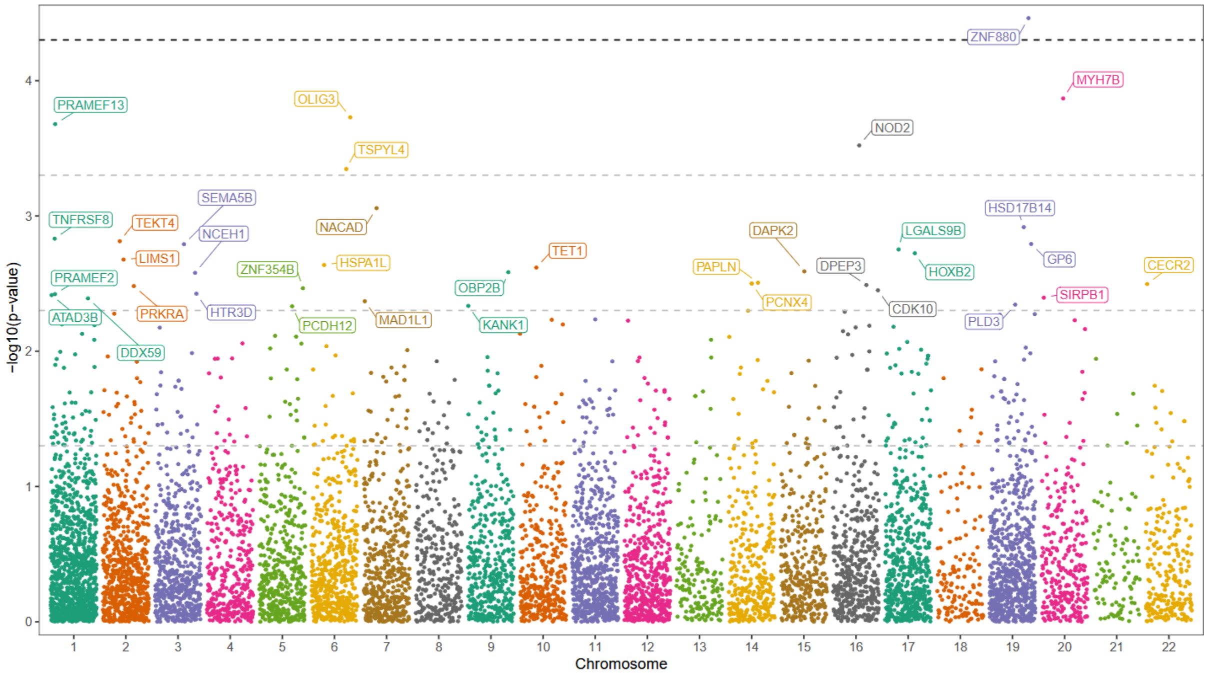


**Suppl. Figure 2: Results from gene-level analysis with MAGMA for all genes**

Horizontal dashed lines represent p-value thresholds of $5x10^{-5}$, $5x10^{-4}$, $5x10^{-3}$, and 0.05 (nominal significance), respectively, considered from top to bottom.

No gene-level association surpassed the significance threshold after multiple-testing correction considering all genes ($2.5x10^{-6}$, not depicted on the graph).

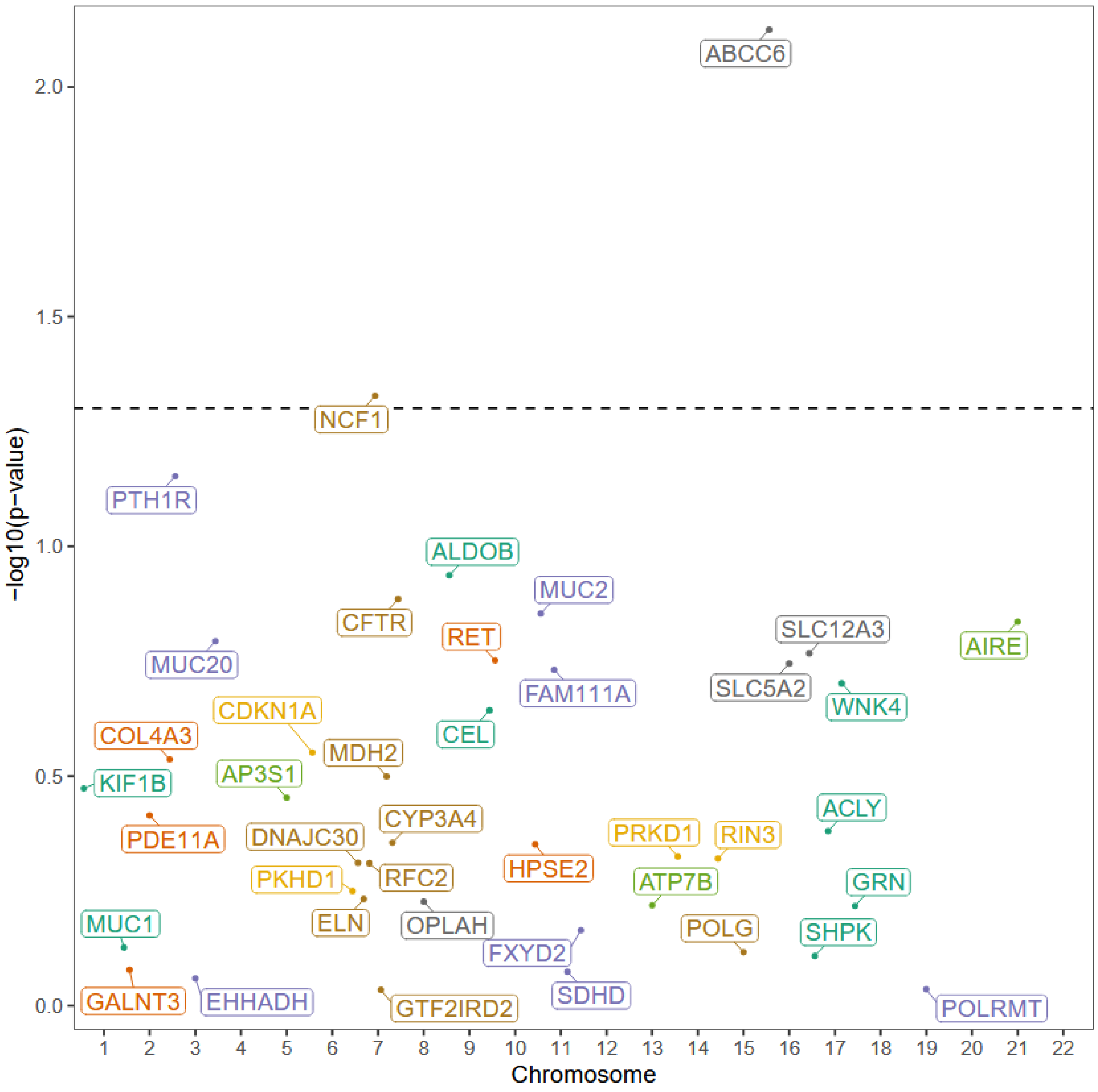


**Suppl. Figure 3: Results from gene-level analysis with MAGMA for novel, i.e., non-established genes nominated by Exomiser**

The horizontal dashed line represents p-value threshold of 0.05.

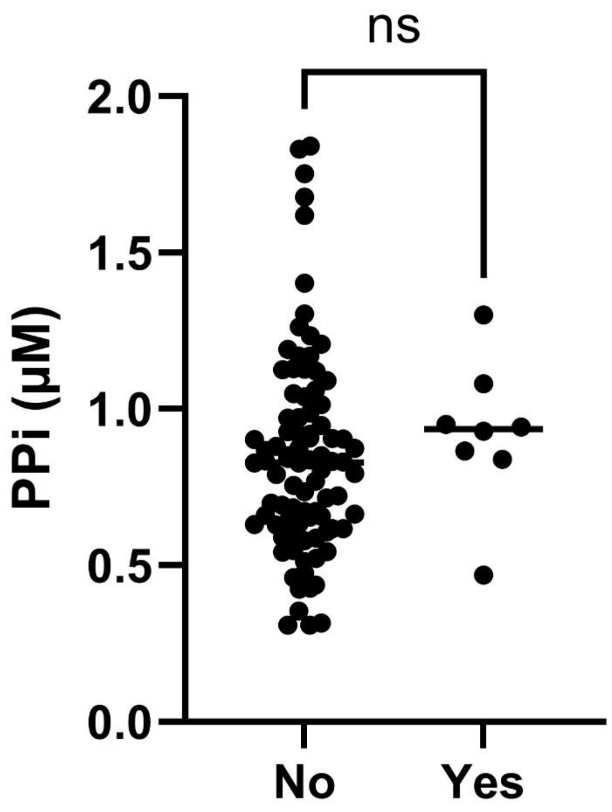


**Suppl. Figure 4: Inorganic plasma pyrophosphate levels in heterozygous carriers of LP/P *ABCC6* variants** with ("Yes") and without ("No") a history of kidney stones. No significant difference is observed between individuals with and without a history of kidney stones ($p$ = 0.773, unpaired t-test (Welch)).
